# Outcomes of Total Neoadjuvant Therapy for Locally Advanced Rectal Cancer: A Prospective Observational Real-World Cohort in Johannesburg, South Africa

**DOI:** 10.64898/2026.10.09.26365104

**Authors:** Saabira Amod, Mariza Tunmer Boshoff, Lisa Campbell-Roper, Georgia Demetriou, Petra Gaylard, Tzipora Krawitz, Deirdre Kruger, June Fabian, Nadine Harran

## Abstract

**Background:** Total neoadjuvant therapy (TNT) is increasingly used for treatment of locally advanced rectal cancer (LARC) to improve treatment compliance, oncological outcomes and opportunities for organ preservation. However, evidence describing its implementation and outcomes in low- and middle-income countries remains limited. This study evaluated the feasibility, treatment delivery, oncological outcomes and organ preservation outcomes of TNT in a real-world South African setting.

**Methods:** This was a retrospective review of a prospective cohort of adults with histologically confirmed, non-metastatic LARC treated with TNT at the Wits Donald Gordon Medical Centre, Johannesburg, between March 2022 and January 2026. Treatment was individualised following multidisciplinary review and included induction and/or consolidation systemic chemotherapy with either long- or short-course radiotherapy. Following TNT, treatment response was assessed clinically, endoscopically and radiologically. Patients with a clinical complete response (cCR) were offered watch-and-wait (WW) surveillance, while those with an incomplete response were recommended for total mesorectal excision (TME). The primary outcome was organ preservation, assessed as TME-free survival. Secondary outcomes included tumour regrowth, disease-free survival (DFS), overall survival (OS), and local recurrence-free survival (LRFS) following TME.

**Results:** Forty-six patients were included; median age was 63 years, 67.4% were male, 82.6% had T3-T4 tumours and 80.4% had node-positive disease. TNT was completed as planned in 95.7% of patients. Grade ≥3 toxicity occurred in 19.5% during chemotherapy and 8.9% during radiotherapy. Twenty-one patients (45.7%) achieved a cCR and entered WW surveillance. TME-free survival was 46% (95% CI 31-60) at 1-year and 18% (95% CI 8-32) at 3-years. Three-year OS and DFS were 87% (95% CI 64-96) and 76% (95% CI 53-89), respectively. Among patients managed with WW, 3-year regrowth-free survival was 53% (95% CI 26-75). Three-year LRFS following TME was 94% (95% CI 68-99), and all patients undergoing TME or completed salvage surgery achieved an R0 resection.

**Conclusion:** TNT was feasible and well tolerated in this real-world South African cohort, with high treatment completion and favourable medium-term oncological outcomes. Although almost half of patients initially achieved a cCR, declining TME-free and regrowth-free survival demonstrated that durable organ preservation was less frequent, highlighting the importance of structured surveillance and timely access to salvage surgery. These findings support the feasibility of delivering contemporary multimodal rectal cancer care in an appropriately resourced African setting.

## Introduction

Globally, the management of locally advanced rectal cancer (LARC) has evolved substantially over the past two decades, particularly for tumours of the mid and lower rectum. Preoperative radiotherapy or chemoradiotherapy followed by total mesorectal excision (TME), with selective use of adjuvant chemotherapy, has long been the standard treatment approach for LARC. While this approach has resulted in excellent local disease control, it is often at the cost of treatment-related morbidity, including bowel, urinary and sexual dysfunction, as well as the potential need for a permanent stoma, all of which may adversely affect quality of life (1). Recognition of these long-term functional consequences have shifted the treatment priorities of rectal cancer treatment beyond oncological outcomes alone. Consequently, organ preservation (OP) has emerged as an important therapeutic endpoint for patients who achieve a clinical complete response (cCR) following neoadjuvant treatment. In appropriately selected patients, a non-operative “watch-and-wait” (WW) strategy with structured surveillance and timely salvage surgery for local regrowth has demonstrated oncological outcomes comparable to those achieved with immediate surgery, while preserving rectal function and quality of life (2–5).

Total neoadjuvant therapy (TNT) has emerged as an alternative approach to the management of LARC. By delivering all systemic chemotherapy and radiotherapy before surgery, TNT aims to improve compliance with chemotherapy, enhance systemic control through the earlier treatment of occult micrometastatic disease, and increase tumour response rates. These benefits have translated into higher pathological and cCR, facilitating OP in appropriately selected patients on the WW strategy. Landmark randomised clinical trials, including RAPIDO (Rectal cancer And Preoperative Induction therapy followed by Dedicated Operation) and PRODIGE 23 (*Partenariat de Recherche en Oncologie Digestive*), have demonstrated that TNT achieves oncological outcomes that are non-inferior to, and in some settings superior to, those of conventional neoadjuvant strategies, while also increasing cCR rates (6,7). The OPRA (Organ Preservation in patients with Rectal Adenocarcinoma) trial further demonstrated that TNT could facilitate OP through a response-adapted WW strategy without compromising long-term oncological outcomes (5). Collectively, these studies have established TNT as a contemporary standard-of-care option for selected patients with LARC.

However, successful implementation of TNT is resource intensive, requiring coordinated multidisciplinary care, reliable access to high-quality imaging, radiotherapy, chemotherapy, colorectal surgical expertise, and a structured WW protocol (8). Consequently, the generalisability of outcomes reported in the above-mentioned clinical trials, conducted predominantly in high-income countries, remains uncertain in low- and middle-income countries (LMICs) such as South Africa, where resource constraints may influence treatment delivery and outcomes.

South Africa exemplifies this uncertainty. In 2022, the specialist Colorectal Unit (CRU) at the Wits Donald Gordon Medical Centre (WDGMC), a private academic teaching hospital in Johannesburg, South Africa, introduced TNT for patients with mid- and low-rectal LARC. This study aimed to evaluate the feasibility of implementing TNT within this setting and to determine whether clinical, oncological, and OP outcomes were comparable to those reported in international studies.

## Methods

### Study design, setting and participants

This was a retrospective review of a prospective cohort of patients with LARC treated by the CRU at WDGMC between March 2022 and January 2026. Patients were eligible for inclusion if they were 18 years or older, had histologically confirmed rectal adenocarcinoma without evidence of distant metastases, and were treated with TNT with curative intent. Tumours were classified according to the distance from the anal verge as low (< 5cm), mid (5-10cm), or high (> 10cm), with baseline staging based on clinical and radiological assessment. Patients were excluded if they were pregnant, had previous or concomitant second malignancy, had received prior radiotherapy precluding TNT, or had previous chemotherapy or surgery for rectal cancer. Patients with incomplete clinical, radiological, treatment or follow-up data were excluded from the analysis.

### Treatment regimens

Treatment was individualised following multidisciplinary team review according to tumour characteristics, patient factors and treatment intent. TNT comprised radiotherapy and systemic chemotherapy delivered before definitive surgical management or consideration of WW surveillance. Systemic chemotherapy was administered either before radiotherapy (induction chemotherapy), after radiotherapy (consolidation chemotherapy), or both before and after radiotherapy, referred to as sandwich therapy. Commonly used systemic chemotherapy regimens included FOLFOX (folinic acid, fluorouracil [5FU] and oxaliplatin), CAPOX (capecitabine and oxaliplatin), capecitabine only, FOLFOXIRI (folinic acid, 5FU, oxaliplatin and irinotecan), FOLFIRINOX (folinic acid, 5FU, irinotecan and oxaliplatin), 5FU only, and 5FU and leucovorin. Radiotherapy was delivered using either a long-course or short-course schedule. Following completion of the planned neoadjuvant regimen, patients underwent clinical and radiological restaging and were subsequently managed with either TME or WW surveillance according to treatment response.

Post-treatment response was assessed using digital rectal examination, endoscopy, pelvic magnetic resonance imaging (MRI) and computed tomography (CT) of the chest, abdomen and pelvis. Restaging was performed as part of routine care using the first available post-TNT assessment, without restriction to a predefined time interval. Response to treatment was categorised as a clinical complete response (cCR), near-complete clinical response (nCR) or incomplete clinical response (iCR). cCR was defined as the absence of clinical, endoscopic or imaging evidence of residual rectal tumour at post-treatment restaging, whereas iCR were patients who had persistent disease following total neoadjuvant therapy requiring surgical resection. Patients with cCR were offered WW surveillance; those with nCR underwent repeat endoscopic assessment and were reclassified as cCR or iCR; those with iCR were recommended for TME. During WW surveillance, patients with a sustained response continued surveillance, while those with suspected tumour regrowth on clinical, endoscopic or radiological assessment were recommended for salvage surgery. Tumour regrowth was defined as reappearance of tumour in the rectum or regional lymph nodes after an initial cCR and a period of WW surveillance. Regrowth was not classified as locoregional failure if followed by a microscopically negative margin in salvage surgery (R0 resection).

### Data collection

Data were collected using Research Electronic Data Capture (REDCap) tools (9). Variables included demographic and clinical characteristics of age, sex, self-reported ethnicity, body mass index (BMI), family history of cancer, comorbidity burden assessed using the Charlson Comorbidity Index (CCI), tumour characteristics (including date of diagnostic biopsy, distance from the anal verge, baseline tumour and nodal stage, American Joint Committee on Cancer [AJCC] stage), treatment characteristics (TNT sequence, chemotherapy regimen, radiotherapy schedule, treatment interruptions, grade ≥3 toxicities) and outcome data (TNT completion, post-treatment response, operative management, pathological findings, tumour regrowth, recurrence, survival status and date of last follow-up). Tumour, node and metastasis (TNM) staging was used to assign final AJCC stage (10).

### Outcomes

The primary outcome was OP, measured as TME-free survival. TME-free survival was defined as the interval from the date of TNT initiation to date of TME, irrespective of whether surgery was performed following initial response assessment or after tumour regrowth during WW surveillance. For patients with iCR who declined surgery, time was calculated to the date of restaging. For patients who experienced tumour regrowth and either declined surgery or developed unresectable disease, time was calculated to the date of regrowth. Patients who did not undergo TME were censored at the date of last follow-up.

Secondary outcomes included (i) tumour regrowth among patients managed with a WW strategy (ii) disease-free survival (DFS), (iii) overall survival (OS), and (iv) local recurrence-free survival (LRFS) following TME. Tumour regrowth was defined as the interval from treatment initiation to documented regrowth among patients managed with WW. DFS was defined as the interval from treatment initiation to the first occurrence of either locoregional failure, distant metastasis, new invasive colorectal primary cancer, or death from any cause. Locoregional failure was defined as a persistent unresectable primary rectal tumour following neoadjuvant treatment, a microscopically (R1) or macroscopically (R2) positive resection margin of the primary rectal tumour, or recurrence within the primary tumour bed following an R0 resection. OS was defined as the interval from treatment initiation to death from any cause, with surviving patients censored at last follow-up. LRFS following TME was defined as the interval from the date of surgery to locoregional failure. Patients without locoregional failure were censored at last follow-up date.

### Statistical analysis

Baseline characteristics were summarised descriptively. Continuous variables were reported as means with standard deviations if normally distributed and medians with interquartile ranges if non-normally distributed, and categorical variables as frequencies and percentages. TNT completion, toxicity and clinical response rates were reported descriptively as binary outcome variables. Time-to-event outcomes were estimated using Kaplan-Meier methods. Survival estimates were calculated from the date of first treatment unless otherwise specified. One- and three-year estimates were reported where follow-up permitted, with 95% confidence intervals where estimable. OS was compared between patients who underwent TME and those who did not require surgery, and DFS was compared between patients who underwent TME after initial restaging versus after tumour regrowth (calculated from date of surgery), using the log-rank test. Given the cohort size and limited number of events, subgroup comparisons were interpreted cautiously. All analyses were performed using SAS version 9.4, with statistical significance set at p<0.05.

### Ethics, institutional approvals and reporting guidelines

Ethics approval was obtained from the University of the Witwatersrand Human Research Ethics Committee (Medical) (clearance number: M230833). The study is reported in compliance with the STROBE guidelines (11).

## Results

A total of 46 patients met the study inclusion criteria, had histologically confirmed rectal adenocarcinoma, and received TNT with curative intent at WDGMC between March 2022 and January 2026. Overall, the patient cohort had a median (interquartile range, IQR) age of 63 years (IQR 36-79), was predominantly male (n=31, 67.4%), of White ethnicity (n=32, 69.6%), and predominantly overweight, with a median BMI of 27 kg/m^2^ (IQR 23-31); Table 1). Detailed tumour characteristics are summarised in **Table 1** and show that a family history of cancer in a first-degree relative was reported by 65.2% of patients. The median CCI was 4 (range 2-8), indicating a moderate to high comorbidity burden. Overall, 82.6% of patients had T3-T4 tumours and 80.4% had nodal involvement. Consequently, the majority of patients were classified as AJCC stage III (78.3%).

**Table 1.**
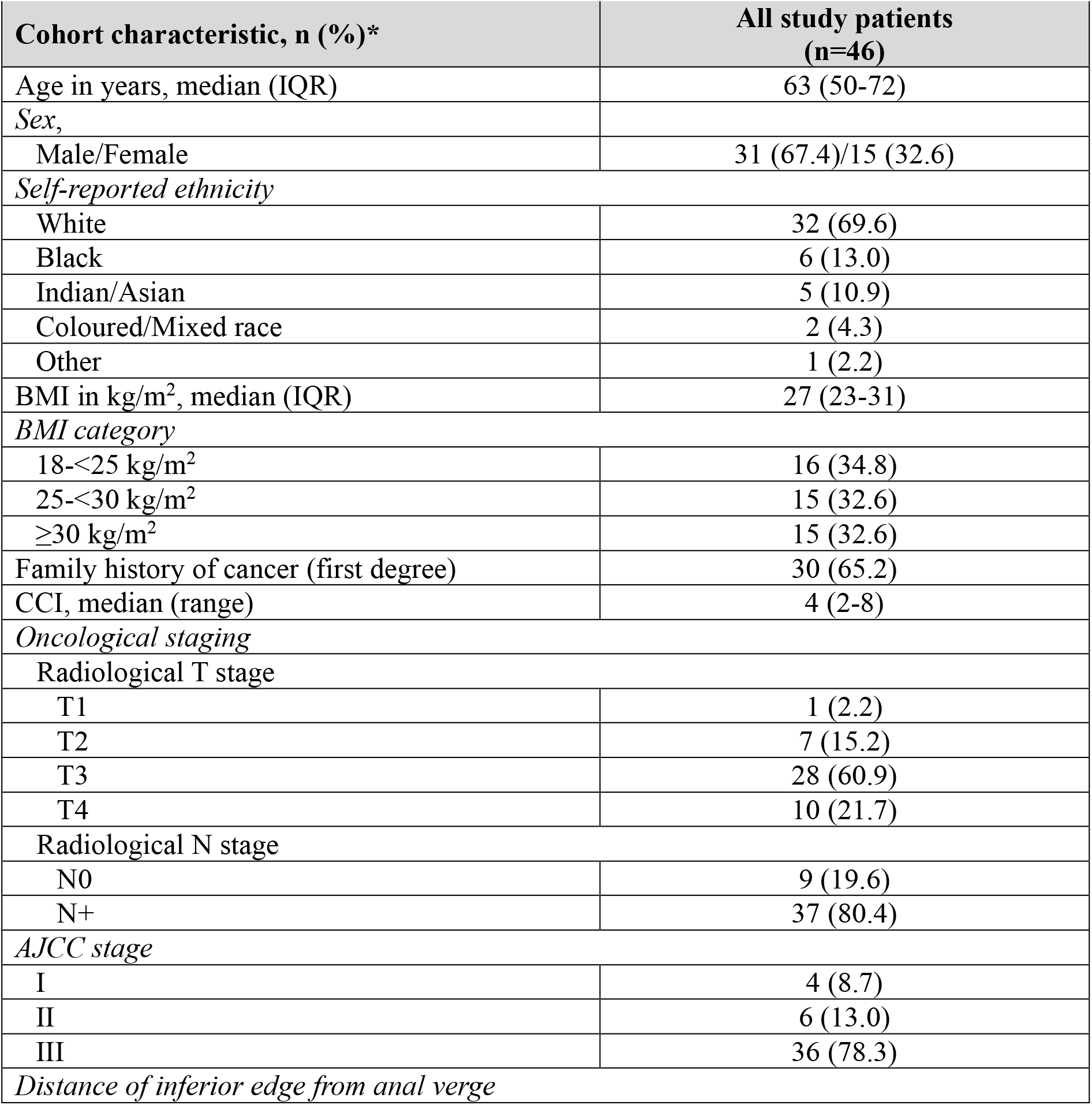
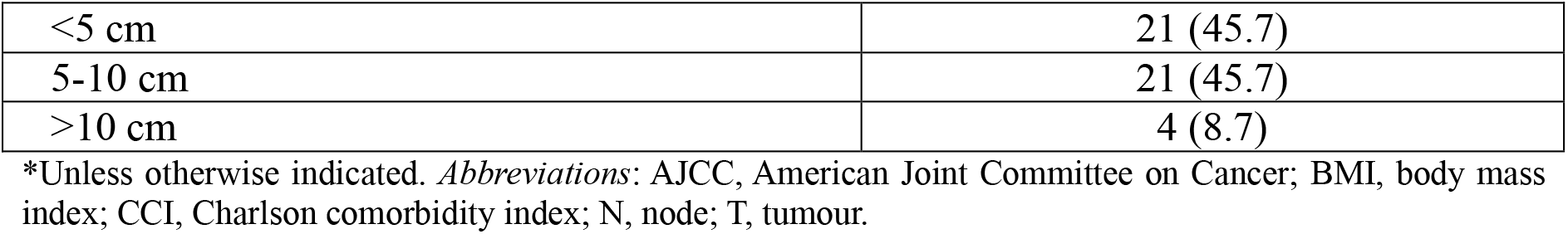
Baseline demographics and tumour characteristics of the study cohort.

### Treatment Regimens

Table 2 describes the treatment regimens for the study cohort. TNT was delivered predominantly using long-course radiotherapy followed by consolidation chemotherapy. Treatment completion was highly achievable irrespective of the choice of treatment regimen, with 95.7% of patients successfully completing their planned TNT regimen. For patients with treatment interruptions, these occurred more frequently during chemotherapy (31.7%) than radiotherapy (10.9%) and were most often related to toxicity. Chemotherapy-related challenges were also reflected in the higher number of Grade ≥3 toxicities reported with chemotherapy (19.5%) compared to radiotherapy (8.9%). The median duration of completed TNT treatment was 201 days (IQR 140-231).

**Table 2.**
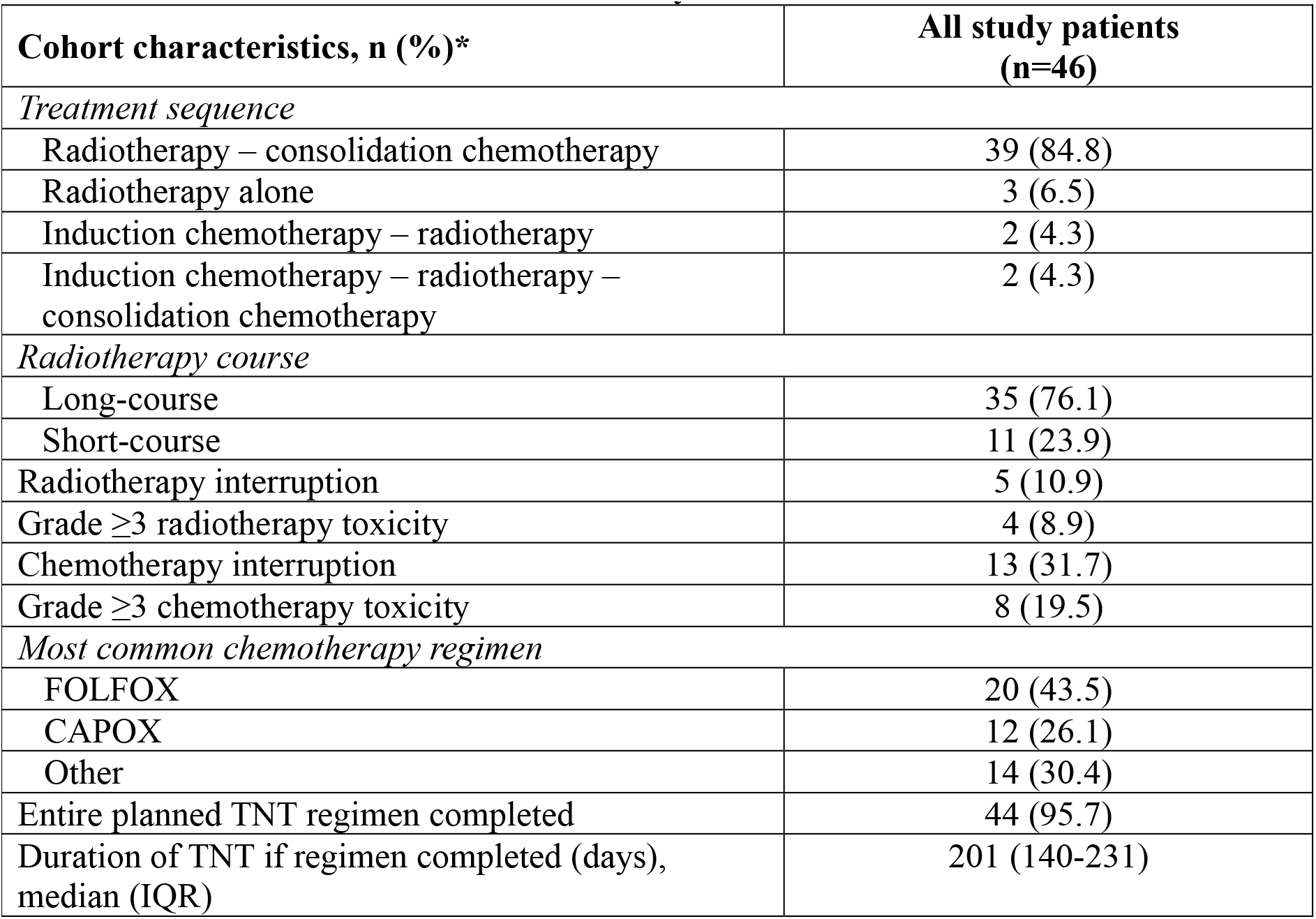
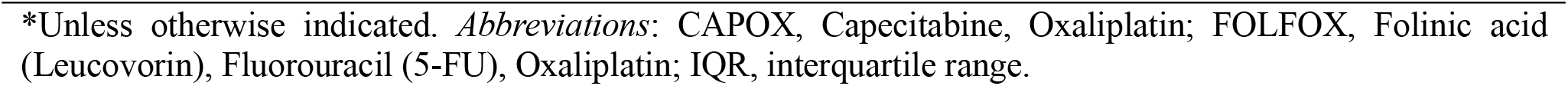
Treatment characteristics of the study cohort.

| Cohort characteristics, n (%)* | All study patients (n=46) |
| --- | --- |
| *Treatment sequence* |  |
| Radiotherapy – consolidation chemotherapy | 39 (84.8) |
| Radiotherapy alone | 3 (6.5) |
| Induction chemotherapy – radiotherapy | 2 (4.3) |
| Induction chemotherapy – radiotherapy – consolidation chemotherapy | 2 (4.3) |
| *Radiotherapy course* |  |
| Long-course | 35 (76.1) |
| Short-course | 11 (23.9) |
| Radiotherapy interruption | 5 (10.9) |
| Grade $\geq 3$ radiotherapy toxicity | 4 (8.9) |
| Chemotherapy interruption | 13 (31.7) |
| Grade $\geq 3$ chemotherapy toxicity | 8 (19.5) |
| *Most common chemotherapy regimen* |  |
| FOLFOX | 20 (43.5) |
| CAPOX | 12 (26.1) |
| Other | 14 (30.4) |
| Entire planned TNT regimen completed | 44 (95.7) |
| Duration of TNT if regimen completed (days), median (IQR) | 201 (140-231) |

\*Unless otherwise indicated. *Abbreviations:* CAPOX, Capecitabine, Oxaliplatin; FOLFOX, Folinic acid (Leucovorin), Fluorouracil (5-FU), Oxaliplatin; IQR, interquartile range.

The median interval from completion of TNT to first restaging was 6 weeks (IQR 3-11 weeks), although timing varied substantially, reflecting real-world clinical practice. Twenty-one patients (45.7%) achieved a cCR and entered the WW surveillance protocol. The remaining 25 patients (54.3%) had an iCR and were recommended for TME. Of these, four declined, two were unfit for surgery, 10 patients had a low anterior resection, and nine had an abdominoperineal resection (APR). A laparoscopic approach was used in 12 patients (63.2%), with the remaining procedures performed either robotically or with an open approach. Irrespective of the surgical approach, an R0 resection was achieved in all patients. Patients who declined or were unfit for surgery were removed from WW surveillance and treated palliatively.

### Outcomes

Table 3 shows 1- and 3-year OP as the primary endpoint, assessed using TME-free survival, and oncological outcomes at 1- and 3-years post TNT, whereas Figure 1 shows the Kaplan-Meier estimate of OP (TME-free survival). TME-free survival decreased over time, with a median TME-free survival rate of 46% (95% CI 31-60%) at 1-year declining to 18% (95%CI 8-32%) at 3-years.

**Table 3:** Organ preservation and oncological outcomes at 1-and 3-years post total neoadjuvant therapy (TNT)

| Outcome parameters | 1-year follow-up<br>% (95% CI) | 3-year follow-up<br>% (95% CI) |
| --- | --- | --- |
| **Primary outcome (organ preservation)** |  |  |
| • TME-free survival | 46 (31-60) | 18 (8-32) |
| **Secondary outcomes** |  |  |
| • Overall survival | 100* | 87 (64-96) |
| • Disease-free survival | 98 (84-99) | 76 (53-89) |
| • Regrowth-free survival, watch-and-wait patients | 89 (62-97) | 53 (26-75) |
| • Local recurrence-free survival after TME | 100* | 94 (68-99) |

\* NE, not estimable (no events by this time point). All times calculated from date of treatment initiation. Six DFS events occurred during follow-up: three deaths, two distant metastases, one locoregional failure *Abbreviations:* CI, confidence interval; TME, total mesorectal excision.

**Figure 1:**
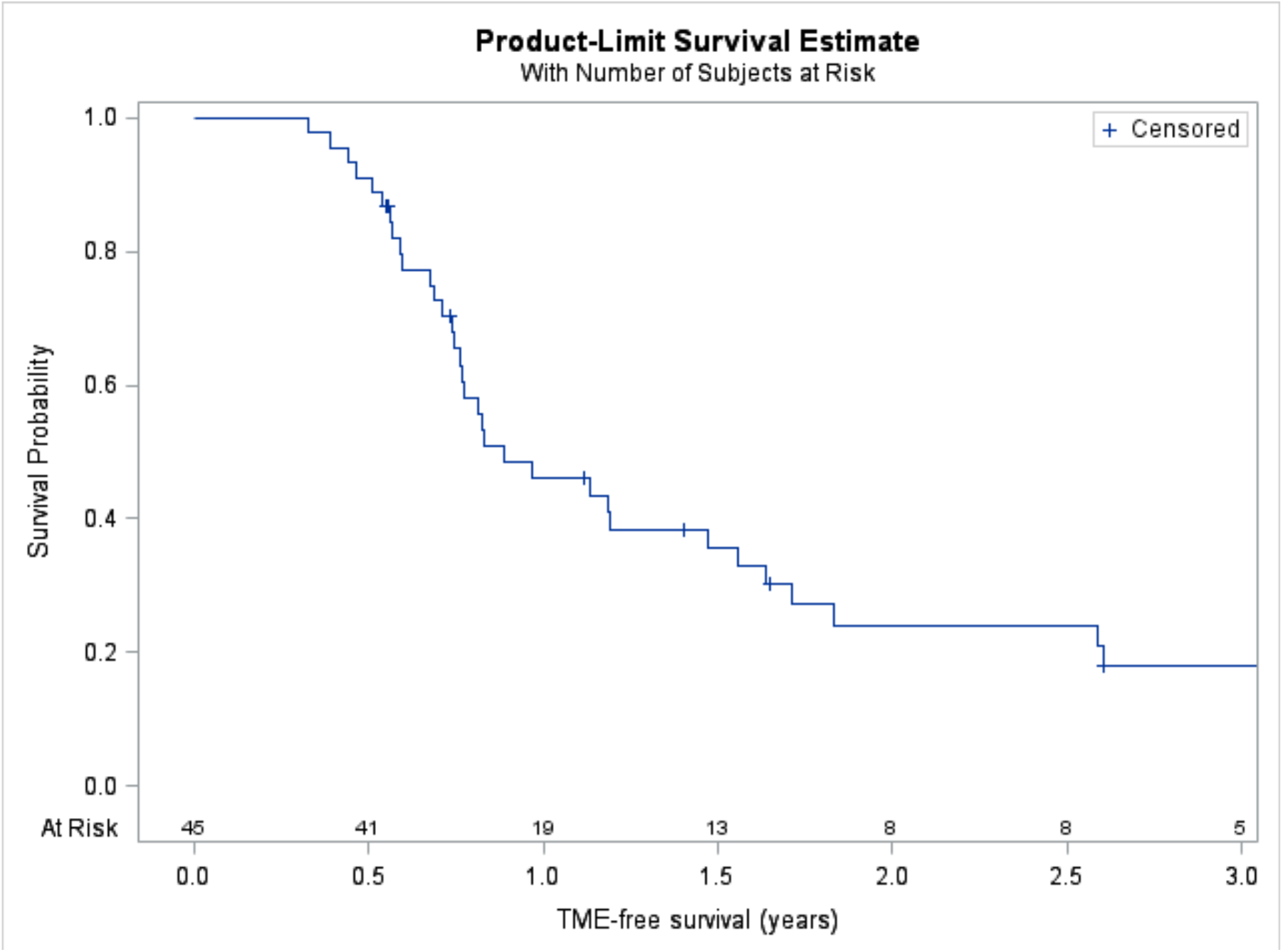
Kaplan-Meier estimate of TME-free survival (OP) for the study cohort

No deaths were observed during the first year post-TNT and an estimated 3-year overall survival of 87% (95% CI 64-96%) was observed. Disease-free survival was high at 98% (95% CI 84-99%) at 1-year and was maintained at 76% (95% CI 53-89%) at 3-years, demonstrating favourable medium-term oncological outcomes. Among patients on WW surveillance, most local regrowth events occurred beyond the first year of surveillance, yet local recurrence-free outcomes following TME were favourable at 94% (95% CI 68-99%) after 3-years. This is demonstrated in Figure 2.

**Figure 2:**
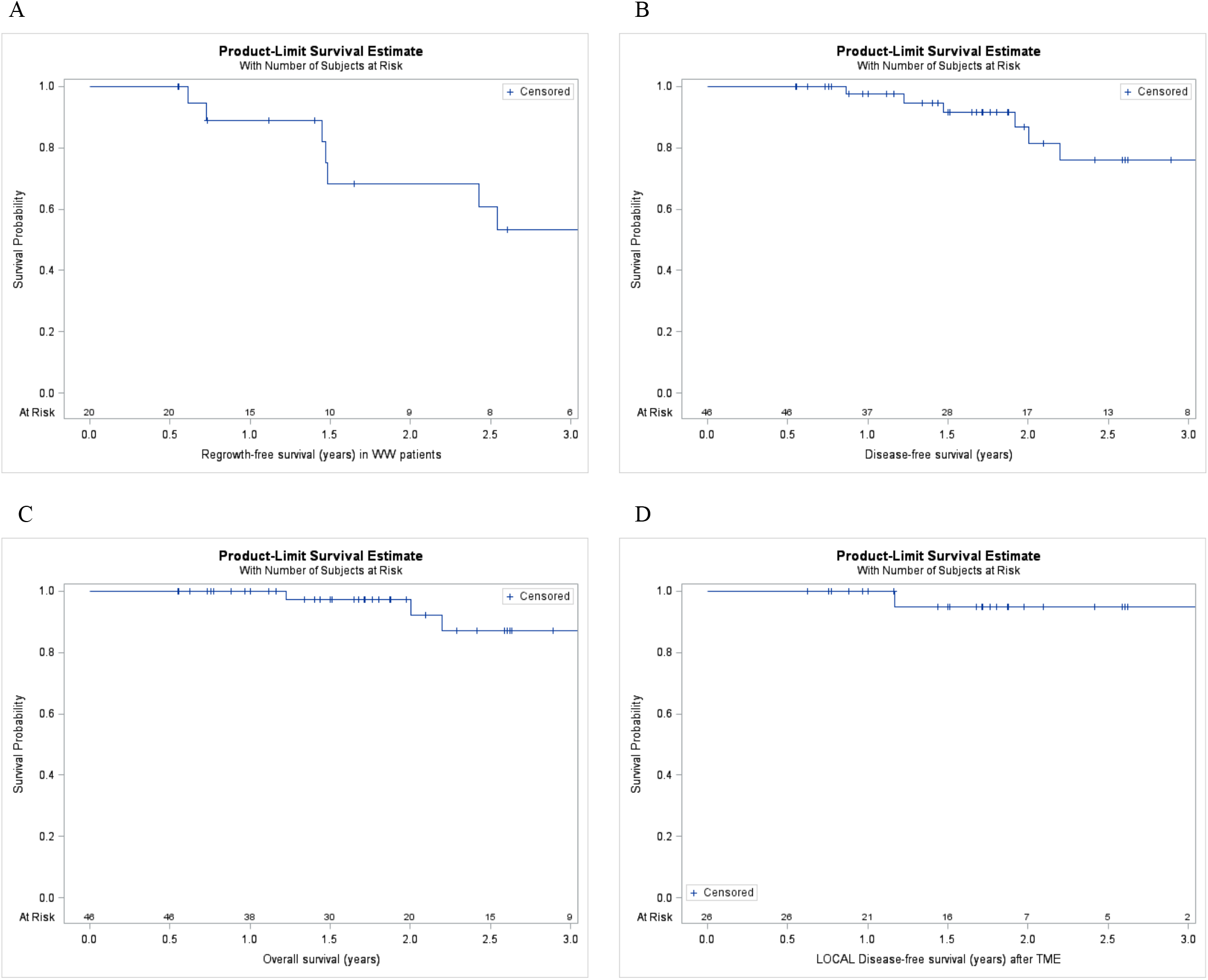
Kaplan-Meier plots of **A**: regrowth-free survival in watch-and-wait patients; **B**: disease-free survival; **C**: overall survival; and **D**: local recurrence-free survival

Of the 21 patients (45.7%) managed with WW surveillance, tumour regrowth occurred in 10 patients (47.6%). One patient had chemotherapy and one patient received no further treatment. Salvage surgery was recommended for the remaining eight patients and had been performed in seven at the time of analysis; the eighth patient was scheduled for an APR. Of those who had surgery, one had a low anterior resection and six had an APR, with an R0 resection achieved in all seven patients.

## Discussion

In this prospective, observational cohort of patients with LARC, TNT was feasible to deliver with high treatment completion rates and favourable oncological outcomes. Overall, 95.7% of patients completed their planned TNT regimen, 45.7% achieved a cCR and entered WW surveillance, and estimated 3-year OS and DFS were 87% and 76%, respectively. However, sustained OP was less frequent, with TME-free survival declining from 46% at 1-year to 18% at 3-years and regrowth-free survival among WW patients decreasing from 89% % at 1-year to 53% at 3-years.

Phenotypically, the reported cohort was similar to local and international studies (5,12). However, this study was conducted in a private specialist setting limiting the representation of the broader South African demographic, as predominantly White patients were assessed. Patients presented with advanced disease comparable to major TNT trials, which largely enrolled patients with stage II-III or high-risk LARC (7,12,13).

Comparable with the OPRA and PRODIGE-23 studies, the completion of radiotherapy was higher than that of systemic chemotherapy, which did not affect treatment completion. (7,13). Severe toxicity rates were lower than those reported in several prospective TNT trials, although direct comparison is limited by differences in treatment regimens, toxicity definitions and reporting methods (13). However, the combination of high treatment completion uncompromised by interruptions with relatively low recorded severe toxicity (grade 3 and higher) supports the tolerability of TNT in routine clinical practice. This supports the feasibility of delivering complex multimodal rectal cancer treatment in a specialist LMIC setting.

The oncological outcomes were favourable and broadly comparable with international studies. Three-year DFS was 76%, closely resembling the approximately 73-76% reported in OPRA, CAO/ARO/AIO-12 and PRODIGE-23 (5,7,12). Three-year OS was 87%, while local recurrence-free survival following TME was 94%. All patients undergoing TME achieved an R0 resection, further supporting effective local disease control in this cohort. Although OP was achieved in a substantial proportion, with 45.7% of patients initially achieving a cCR and entering WW surveillance, TME-free survival declined to 18% by three years. Among the 21 patients managed initially with WW, 10 developed tumour regrowth, with 3-year regrowth-free survival of 53%. These results are less favourable than those reported in OPRA, where 3-year TME-free survival remained substantially higher, particularly following consolidation chemotherapy (53%) (5). The difference may reflect the small sample size, variation in timing of restaging, and differences in response assessment. Importantly, salvage surgery remained effective in that all patients who achieved an R0 resection. These findings reinforce the importance of structured and continued surveillance, as well as timely salvage surgery in patients managed non-operatively on WW.

The study has the following limitations. It was a single-centre observational study with a small sample size and relatively few survival events, limiting statistical precision and subgroup analyses. Treatment regimens and the timing of restaging were not fully standardised, reflecting real-world practice but potentially influencing response assessment and OP outcomes. The cohort was also drawn from a private specialist centre and was predominantly White, limiting generalisability to the broader South African population. These findings are therefore most generalisable to similarly resourced specialist centres in LMICs rather than to all LMIC healthcare settings. Access to high-quality imaging, radiotherapy, chemotherapy, colorectal surgery and structured surveillance may be substantially more limited elsewhere. However, the high-treatment completion rates and favourable oncological outcomes demonstrate that TNT can be implemented successfully in an appropriately resourced African setting.

In conclusion, TNT was feasible and well tolerated in this South African real-world cohort and achieved oncological outcomes broadly comparable with major international studies. However, declining TME-free and regrowth-free survival highlight that durable OP requires careful patient selection, structured surveillance and timely access to salvage surgery. Larger multicentre studies with longer follow-up are needed to assess whether these findings can be reproduced across more diverse LMIC populations.

## Data Availability

All data produced in the present study are available upon reasonable request to the authors

